# Ranking the relative importance of COVID-19 immunisation strategies: a survey of expert stakeholders in Canada

**DOI:** 10.1101/2020.09.16.20196295

**Authors:** Linlu Zhao, Shainoor J. Ismail, Matthew C. Tunis

## Abstract

**Background:** In the face of anticipated limited COVID-19 vaccine supply necessitating the vaccination of certain groups earlier than others, the assessment of values and preferences of stakeholders is an important component of an ethically sound vaccine prioritisation framework.

**Objective:** To establish a preliminary expert stakeholder perspective on the relative importance of pandemic immunisation strategies for different COVID-19 pandemic scenarios at the time of initial COVID-19 vaccine availability.

**Methods:** A survey was conducted by an email process from July 22 to August 14, 2020. Stakeholders included clinical and public health expert groups, provincial and territorial committees and national Indigenous groups, patient and community advocacy representatives and experts, health professional associations, and federal government departments in Canada. Survey results were analysed using descriptive statistics.

**Results:** Of 156 stakeholders contacted, 74 surveys were completed for a participation rate of 47.4%. During an anticipated period of initial vaccine scarcity for all pandemic scenarios, stakeholders generally considered the most important immunisation strategy to be protecting those who are most vulnerable to severe illness and death from COVID-19. This was followed in importance by the strategies to protect healthcare capacity, and to minimise transmission of COVID-19. In this supply constrained context, an immunisation strategy to protect critical infrastructure was considered the least important.

**Conclusion:** The findings of this study provide a timely, preliminary Canadian expert perspective on priority COVID-19 pandemic immunisation strategies to guide early public health planning for an eventual COVID-19 immunisation program. These results fill a gap in the literature and could help advisory groups around the world in their assessment of values and preferences for ethical guidelines for COVID-19 vaccine allocation.

## INTRODUCTION

Global efforts are underway to develop a novel coronavirus disease 2019 (COVID-19) vaccine and work is progressing at an unprecedented pace [1]. Once a successful COVID-19 vaccine becomes available, the initial supply is not expected to be sufficient to immunise the entire population right away. Certain groups will likely receive the vaccine earlier than others.

Many countries have started working on COVID-19 vaccine prioritisation strategies via their National Immunization Technical Advisory Groups (NITAGs). Notably, the United Kingdom’s Joint Committee on Vaccination and Immunisation published in June 2020 their interim prioritisation advice, which includes an early emphasis on frontline healthcare workers and those at increased risk of serious disease and death [2]. In Canada, the National Advisory Committee on Immunization (NACI) is identified in the pandemic strategy as the authoritative body for advice on vaccine prioritisation and program design [3].

The goal of Canada’s pandemic response is to minimise serious illness and overall deaths while minimising societal disruption as a result of the COVID-19 pandemic [4]. While all immunisation strategies are important, limited initial vaccine supply will likely necessitate the prioritisation of immunisation strategies to best achieve the pandemic response goal. Immunisation strategies proposed by the NACI Secretariat based on the Canadian Pandemic Influenza Preparedness guidance [3] and with input from NACI’s High Consequence Infectious Disease (HCID) Working Group included the following:

- Protect those who are most vulnerable to severe illness and death from COVID-19;
- Minimise transmission of COVID-19;
- Protect healthcare capacity; and
- Protect critical infrastructure.

Pandemic immunisation strategies need to be established early in order to inform federal, provincial, and territorial vaccine program planning, including which population groups to include in initial vaccination. However, the final pandemic vaccine recommendations in Canada cannot be made until more is known about the pandemic vaccine characteristics (e.g., efficacy, safety, dosing schedule), how well the vaccine works in different populations (e.g., elderly, those with high-risk medical conditions), and the supply situation. Until then, planning for a COVID-19 vaccine program with a clear vision of the relative importance of pandemic immunisation strategies is necessary.

The systematic consideration of ethics, in addition to other factors in vaccine program recommendations such as equity, acceptability, and feasibility, is enabled through evidence-informed tools and an overall framework that NACI uses when developing recommendations [5]. For guidance to uphold the ethical principles of inclusiveness as well as respect for persons and communities, the engagement of stakeholders and assessment of their values and preferences is critical. In addition to this stakeholder survey, NACI will consider survey data [6-8] on the acceptability of COVID-19 vaccines and prioritisation of immunisation strategies in the general public and high-risk groups in order to inform its guidance.

As part of the planning for pandemic influenza, a 2006 study of university students and staff investigated values in the allocation of scarce resources [9]. The preferred priority was to save the most lives (39.9%), and while 22.4% preferred a ranking system, 20.4% of respondents would save those most likely to die. In that study, respondents ranked “high priority” target groups for vaccination as: healthcare workers (89%), emergency workers (85%), children 2–12 years of age (73.6%), essential workers (60.3), and those who are vulnerable (55.6%). These results differ from a survey of the Canadian population in the midst of the COVID-19 pandemic, where the most commonly identified target groups for priority vaccination include individuals with underlying medical conditions (57%), the elderly (53%), healthcare workers (22%), and frontline/essential workers (18%) [10]. Though the methodologies of these studies are different, the marked differences in results reveal the importance of assessing values and preferences of stakeholders in different contexts.

Canada’s previously established pandemic influenza immunisation strategies [3], which took into account the results from the 2006 study, were not directly applicable to the COVID-19 pandemic due to differences in risk groups (e.g., elderly disproportionately affected), transmission (e.g., higher reproductive number), and impact (e.g., social and economic lockdown). There was a need to identify priority COVID-19 pandemic immunisation strategies in a timely fashion in order to inform public health decision-making and immunisation program planning. Therefore, the objective of this study was to conduct a rapid survey of selected expert stakeholders to establish as comprehensively as possible a preliminary Canadian perspective on the relative importance of pandemic immunisation strategies for different COVID-19 pandemic scenarios at the time of initial COVID-19 vaccine availability.

## MATERIAL AND METHODS

The survey was comprised of five questions that asked the respondent to rank, in order of importance with a rank of “1” being the most important, the four aforementioned COVID-19 pandemic immunisation strategies plus an optional respondent-specified strategy for each of the pandemic scenarios presented in Table 1. These scenarios are also visualised along a hypothetical pandemic curve in Figure 1. The respondent was asked to assume that the COVID-19 vaccine is in limited supply for each scenario and that the COVID-19 vaccine is safe and efficacious for all populations for the purposes of the survey. Other information was not collected.

**Table 1.**
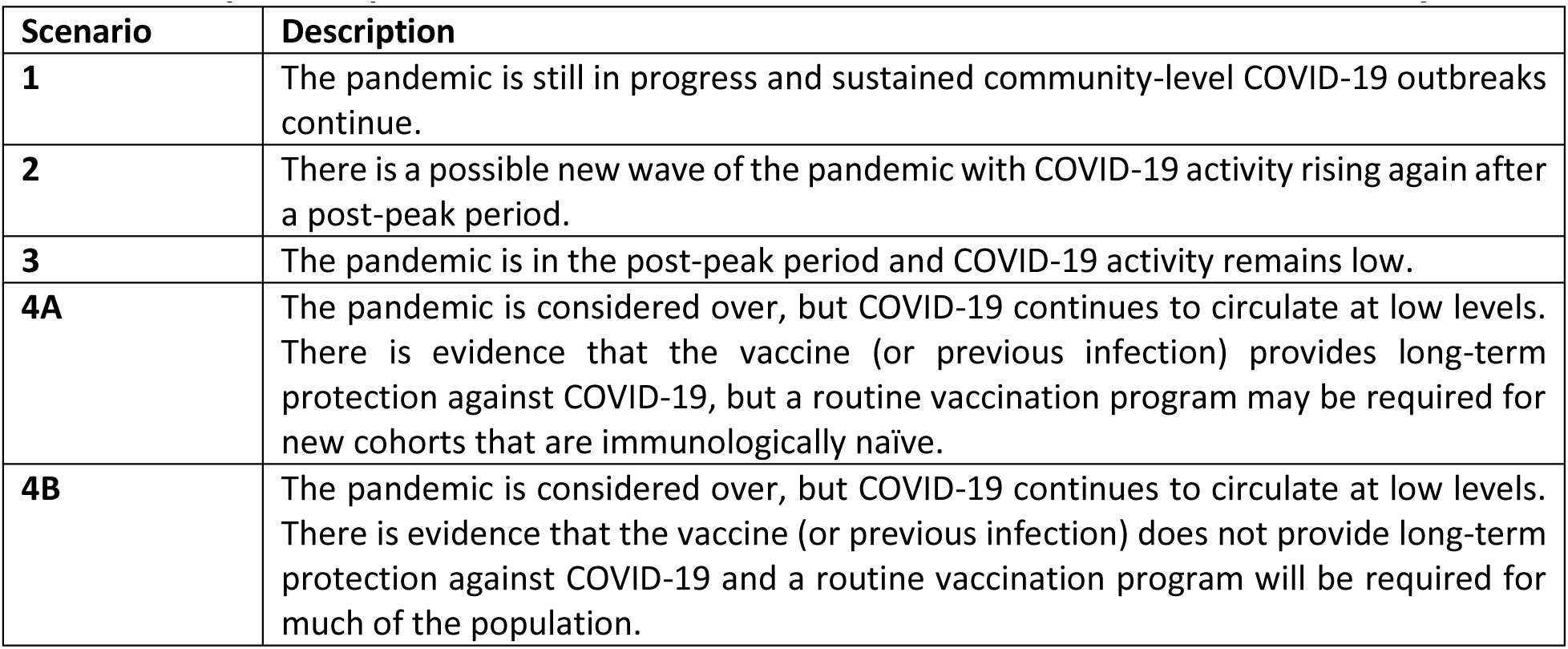
Descriptions of pandemic scenarios at the time of initial COVID-19 vaccine availability.

**Figure 1.**
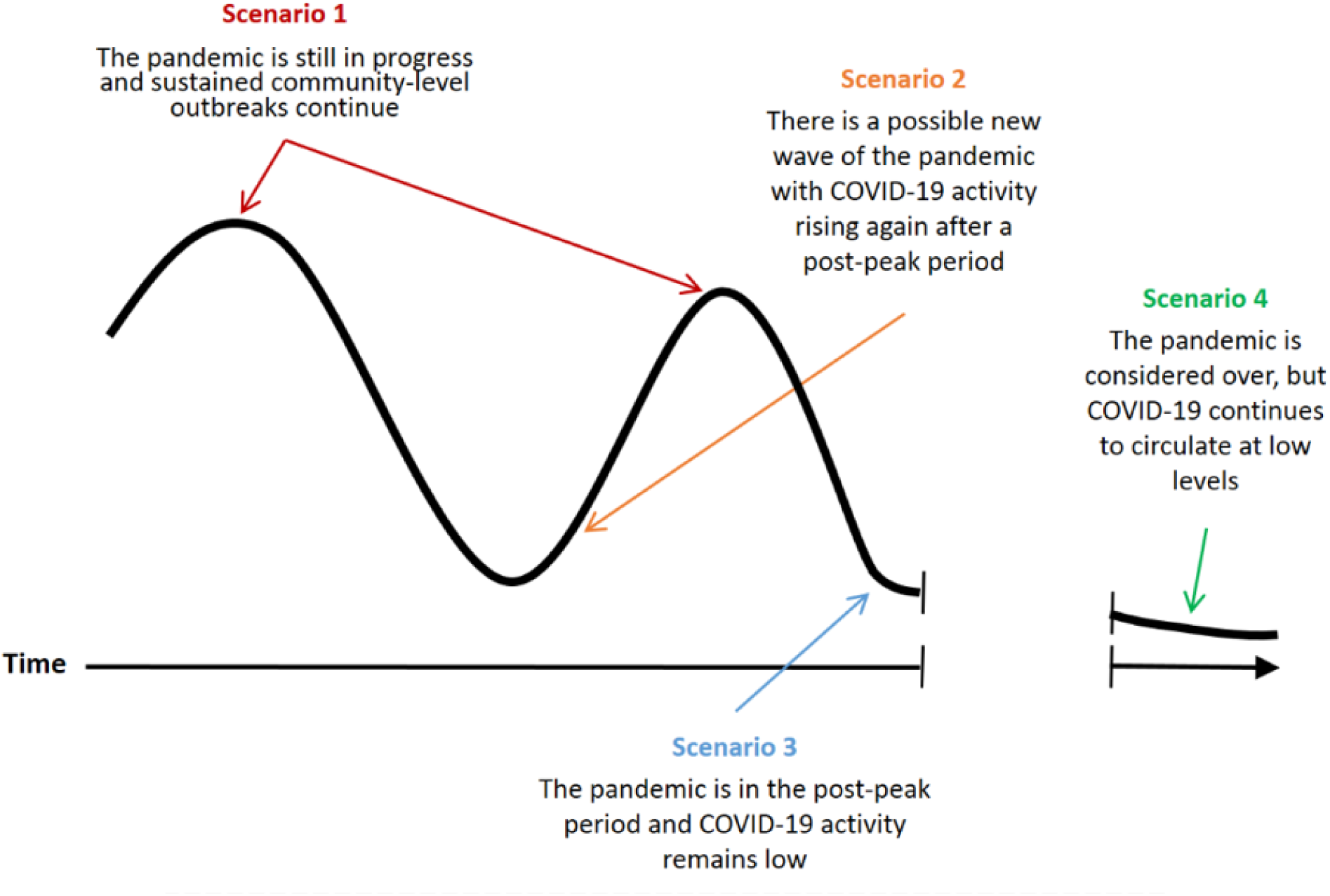
Pandemic scenarios at the time of initial COVID-19 vaccine availability plotted along a hypothetical pandemic curve.

Expert stakeholders were identified through consultations within the Public Health Agency of Canada (PHAC) and with NACI’s HCID Working Group. These stakeholders included members of clinical and public health expert groups involved with PHAC, members of provincial and territorial committees and representatives from national Indigenous groups, patient and community advocacy representatives and experts from the CanCOVID network (https://cancovid.ca/), executives of Canadian health professional associations, and representatives of federal government departments, excluding PHAC (Table 2). An invitation to complete the survey, which was provided as a Word document in English and French, was sent by email to stakeholders in a format that facilitated shared review and discussion within their respective organisations. Members of expert groups (e.g., NACI) each provided individual expert responses, whereas organisational or provincial/territorial representatives each provided a single response on behalf of their organisation or jurisdiction.

**Table 2.**
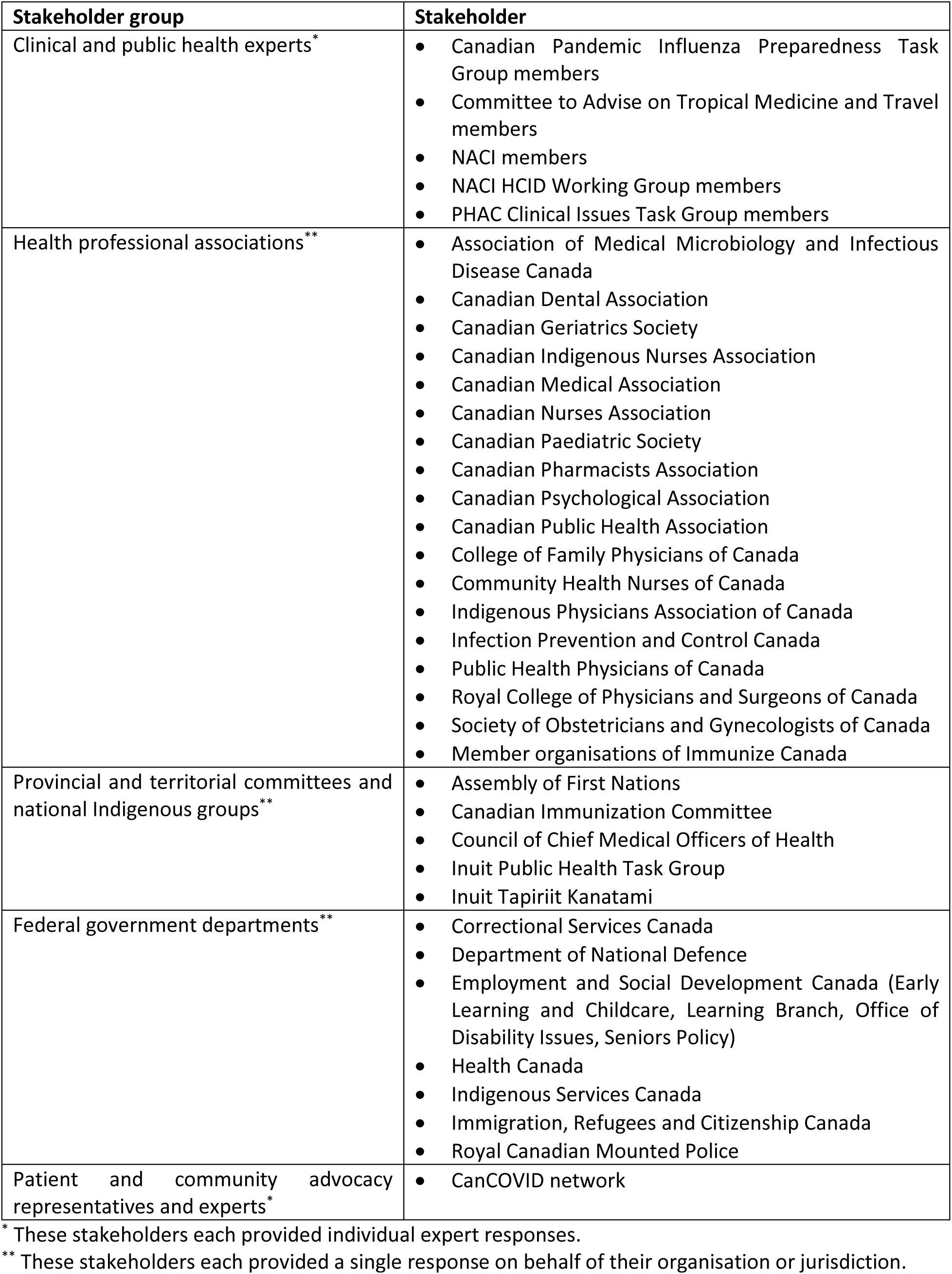
List of surveyed expert stakeholders.

| **Stakeholder group** | **Stakeholder** |
| --- | --- |
| Clinical and public health experts* | • Canadian Pandemic Influenza Preparedness Task Group members<br>• Committee to Advise on Tropical Medicine and Travel members<br>• NACI members<br>• NACI HCID Working Group members<br>• PHAC Clinical Issues Task Group members |
| Health professional associations** | • Association of Medical Microbiology and Infectious Disease Canada<br>• Canadian Dental Association<br>• Canadian Geriatrics Society<br>• Canadian Indigenous Nurses Association<br>• Canadian Medical Association<br>• Canadian Nurses Association<br>• Canadian Paediatric Society<br>• Canadian Pharmacists Association<br>• Canadian Psychological Association<br>• Canadian Public Health Association<br>• College of Family Physicians of Canada<br>• Community Health Nurses of Canada<br>• Indigenous Physicians Association of Canada<br>• Infection Prevention and Control Canada<br>• Public Health Physicians of Canada<br>• Royal College of Physicians and Surgeons of Canada<br>• Society of Obstetricians and Gynecologists of Canada<br>• Member organisations of Immunize Canada |
| Provincial and territorial committees and national Indigenous groups** | • Assembly of First Nations<br>• Canadian Immunization Committee<br>• Council of Chief Medical Officers of Health<br>• Inuit Public Health Task Group<br>• Inuit Tapiriit Kanatami |
| Federal government departments** | • Correctional Services Canada<br>• Department of National Defence<br>• Employment and Social Development Canada (Early Learning and Childcare, Learning Branch, Office of Disability Issues, Seniors Policy)<br>• Health Canada<br>• Indigenous Services Canada<br>• Immigration, Refugees and Citizenship Canada<br>• Royal Canadian Mounted Police |
| Patient and community advocacy representatives and experts* | • CanCOVID network |

\* These stakeholders each provided individual expert responses.
\*\* These stakeholders each provided a single response on behalf of their organisation or jurisdiction.

The survey was conducted between July 22 and August 14, 2020. An email reminder was sent to non-responders to optimise the participation rate. The participation rate was calculated by dividing the number of responders by the sum of responders and non-responders. Survey results were analysed using descriptive statistics across all respondents to identify overall trends and by stakeholder group to assess for any differences in prioritisation among stakeholder groups. Trends in the rankings for each pandemic scenario were assessed by descriptive analysis in two ways: taking the average (mean, median, and mode) ranking and comparing the percentage of each ranking contributing to the total for each COVID-19 pandemic immunisation strategy for different pandemic scenarios at the time of initial COVID-19 vaccine availability.

This study received approval from the Health Canada and the Public Health Agency of Canada Research Ethics Board (REB 2020-011P). The survey invitation letter and the study survey are available in the supplemental materials.

## RESULTS

Of 156 stakeholders contacted, 74 surveys were completed for a participation rate of 47.4%. A total of 22 (29.7%) respondents were members of clinical or public health expert groups involved with PHAC, 19 (25.7%) were patient or community advocacy representatives or experts from the CanCOVID network, 16 (21.6%) were executives of Canadian health professional associations, nine (12.2%) were members of provincial and territorial committees or national Indigenous groups, and eight (10.8%) were representatives of federal government departments. Two respondents returned blank surveys and these were not counted as completed surveys. Two respondents did not complete one of the survey questions and an additional 10 respondents did not provide distinct ranks in the order of importance (i.e., two or more strategies were ranked equivalently) for one (n=4 respondents) or more (n=6 respondents) survey questions. Responses with non-distinct ranks were not included in the analysis (sensitivity analysis including all responses was performed). Ten respondents also ranked strategies out of five, as an “other” strategy was specified, for at least one scenario; these other respondent-specified strategies were all considered by the study investigators to fall under one of the four predetermined strategies, but the rankings out of five were retained for analysis.

For all pandemic scenarios, both descriptive analysis approaches showed that stakeholders generally ranked the strategies in the following order from most to least important:

1. Protect those who are most vulnerable to severe illness and death from COVID-19
2. Protect healthcare capacity
3. Minimise transmission of COVID-19
4. Protect critical infrastructure

In subgroup analysis by stakeholder group, the trends were less clear due to smaller sample sizes, but the strategy to protect those who are most vulnerable to severe illness and death from COVID-19 remained the most important in all stakeholder groups and across pandemic scenarios. Sensitivity analysis including all responses, including those that had non-distinct ranks, did not differ in overall trends.

The average (mean, median, and mode) rankings and stacked bar charts of rankings for COVID-19 pandemic immunisation strategies for different pandemic scenarios at the time of initial COVID-19 vaccine availability are presented in Table 3 and Figure 2, respectively.

**Table 3.** Average (mean, median, and mode) rankings for COVID-19 pandemic immunisation strategies for different pandemic scenarios at the time of initial COVID-19 vaccine availability.

|  | COVID-19 pandemic immunisation strategies |  |  |  |
| --- | --- | --- | --- | --- |
|  | Protect those who are most vulnerable to severe illness and death from COVID-19 | Minimise transmission of COVID-19 disease | Protect critical infrastructure | Protect healthcare capacity |
| **Scenario 1:** The pandemic is still in progress and sustained community-level COVID-19 outbreaks continue. |  |  |  |  |
| Mean | 1.7 | 2.9 | 3.3 | 2.2 |
| Median | 1 | 3 | 4 | 2 |
| Mode | 1 | 4 | 4 | 2 |
| **Scenario 2:** There is a possible new wave of the pandemic with COVID-19 activity rising again after a post-peak period. |  |  |  |  |
| Mean | 1.7 | 2.7 | 3.4 | 2.3 |
| Median | 1 | 3 | 4 | 2 |
| Mode | 1 | 4 | 4 | 2 |
| **Scenario 3:** The pandemic is in the post-peak period and COVID-19 activity remains low. |  |  |  |  |
| Mean | 1.4 | 2.7 | 3.4 | 2.6 |
| Median | 1 | 2 | 4 | 3 |
| Mode | 1 | 2 | 4 | 3 |
| **Scenario 4A:** The pandemic is considered over, but COVID-19 continues to circulate at low levels. There is evidence that the vaccine (or previous infection) provides long-term protection against COVID-19, but a routine vaccination program may be required for new cohorts that are immunologically naïve. |  |  |  |  |
| Mean | 1.3 | 2.9 | 3.4 | 2.6 |
| Median | 1 | 3 | 4 | 3 |
| Mode | 1 | 2 | 4 | 2 |
| **Scenario 4B:** The pandemic is considered over, but COVID-19 continues to circulate at low levels. There is evidence that the vaccine (or previous infection) does not provide long-term protection against COVID-19 and a routine vaccination program will be required for much of the population. |  |  |  |  |
| Mean | 1.6 | 2.8 | 3.4 | 2.3 |
| Median | 1 | 3 | 4 | 2 |
| Mode | 1 | 4 | 4 | 2 |

**Figure 2.**
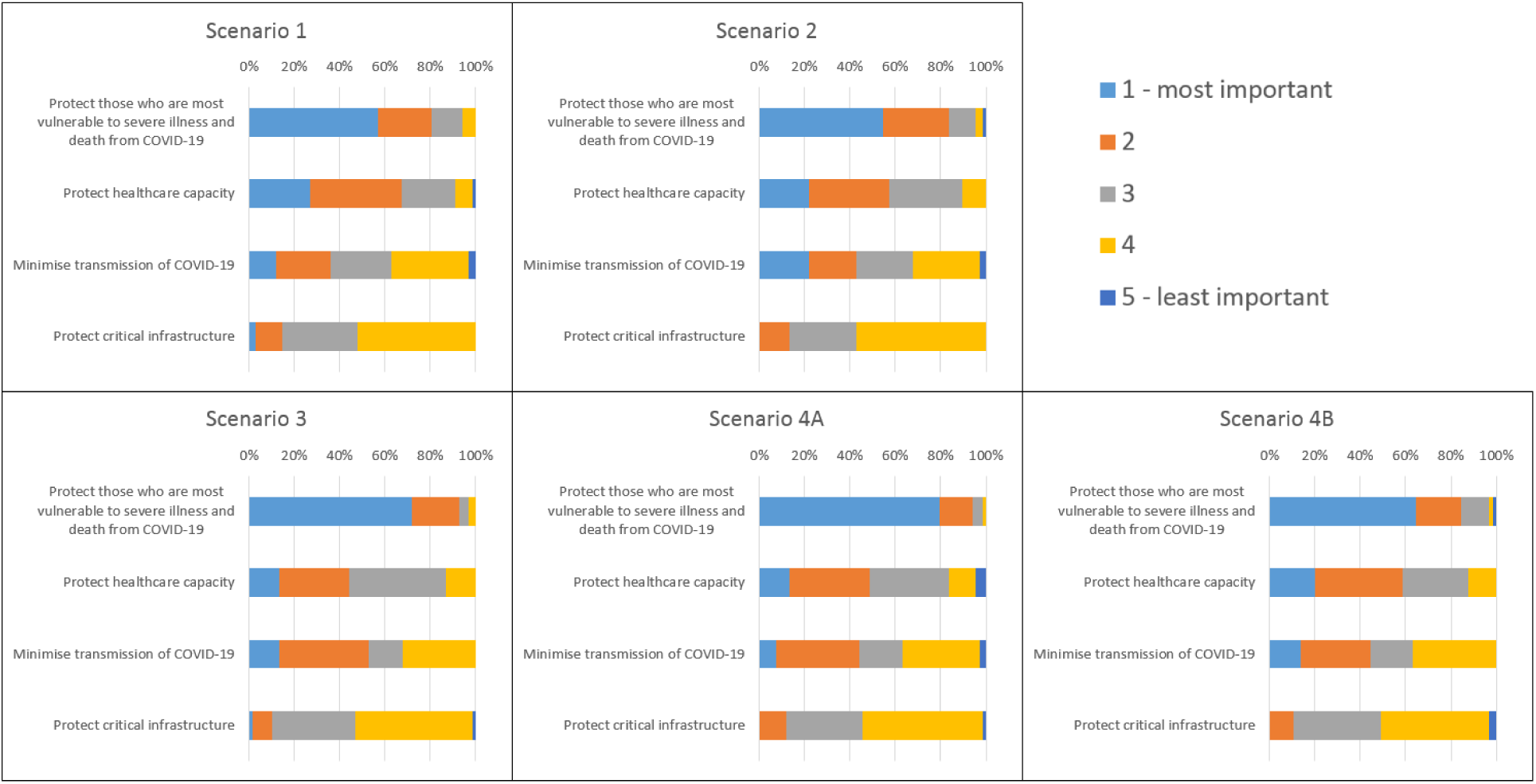
Stacked bar charts comparing the percentage of each ranking contributing to the total for COVID-19 pandemic immunisation strategies for different pandemic scenarios at the time of initial COVID-19 vaccine availability. Scenario 1: The pandemic is still in progress and sustained community-level COVID-19 outbreaks continue. Scenario 2: There is a possible new wave of the pandemic with COVID-19 activity rising again after a post-peak period. Scenario 3: The pandemic is in the post-peak period and COVID-19 activity remains low. Scenario 4A: The pandemic is considered over, but COVID-19 continues to circulate at low levels. There is evidence that the vaccine (or previous infection) provides long-term protection against COVID-19, but a routine vaccination program may be required for new cohorts that are immunologically naïve. Scenario 4B: The pandemic is considered over, but COVID-19 continues to circulate at low levels. There is evidence that the vaccine (or previous infection) does not provide long-term protection against COVID-19 and a routine vaccination program will be required for much of the population.

## DISCUSSION

The present study showed that the surveyed stakeholders generally considered the most important immunisation strategy to be that of protecting those who are most vulnerable to severe illness and death from COVID-19 during the period of initial vaccine scarcity. This was followed in importance by the strategies to protect healthcare capacity and to minimise transmission of COVID-19 disease. In this supply constrained context, an immunisation strategy to protect critical infrastructure was considered the least important.

There are a number of important limitations to consider when interpreting the findings of this study. First, stakeholders were forced to treat the immunisation strategies presented in the survey as distinct, when in reality these strategies are overlapping to some degree. For example, those working in long-term care facilities could be targeted under all four immunisation strategies that were presented for ranking. Second, this study surveyed “key informant” stakeholders who acted as a proxy for their organisation or stakeholder group. Though respondents were encouraged to consult with others in their organisations, the survey responses may not be representative opinions of the respective organisations or groups. Third, the survey questions presented broad concepts that were open to interpretation. Respondents likely made differing assumptions based on their values and preferences in order to provide rankings.

Despite these limitations, the overall ranking of this expert survey mirrored surveys of the general public on the prioritisation of pandemic immunisation strategies. Canada’s COVID-19 Snapshot Monitoring Study (COSMO Canada) is a longitudinal study that surveyed a representative sample of approximately 2000 Canadians from April through September 2020 in eight waves [6]. When asked in Wave 7 (August 13–17, 2020) which immunisation strategies they would prioritise if COVID-19 vaccine supply is limited, a majority of respondents identified protecting those most vulnerable (51%) and protecting healthcare capacity (28%) as the most important strategies to determine which groups should receive the COVID-19 vaccine first when there is not enough vaccine for everyone when it first becomes available [7]. This was followed by minimising transmission (15%) and protecting critical infrastructure (5%). In the 2006 study of University of Alberta students and staff on the allocation of scarce resources during an influenza pandemic, the top choice for a priority access plan to the pandemic vaccine was to save the most lives [9]. This alignment lends confidence to the findings of the present study; however, these preliminary priority pandemic immunisation strategies will need to be further validated in follow up surveys, particularly with the general public and high-risk groups, which are planned for fall 2020.

The findings of this study provide a timely, preliminary expert perspective on priority COVID-19 pandemic immunisation strategies to guide early public health planning for the eventual COVID-19 vaccine or vaccines, including the development of guidance by NACI on the prioritisation of COVID-19 vaccines. The results of this analysis could prove useful for other countries around the world planning allocation of limited initial supplies of COVID-19 vaccine.

## Data Availability

Not applicable.

## ACKNOWLEDGEMENTS

We would like to thank the NACI HCID Working Group (Caroline Quach [Chair], Shelley Deeks [Vice-Chair], Yen Bui, Kathleen Dooling, Robyn Harrison, Kyla Hildebrand, Michelle Murti, Jesse Papenburg, Robert Pless, Nathan Stall, and Stephen Vaughan) for their contributions to this study. We would also like to thank Michelle Matthieu-Higgins and Renee Goddard for their project management support.

## SUPPLEMENTARY MATERIALS

**Supplementary Material 1**.

Sample survey invitation letter in English and French.

**Supplementary Material 2**.

Study survey in English.

**Supplementary Material 3**.

Study survey in French.

